# The Effects of Stress Across the Lifespan on the Brain, Cognition and Mental Health: A UK Biobank study

**DOI:** 10.1101/2021.05.11.21256756

**Authors:** Elizabeth McManus, Hamied Haroon, Niall W. Duncan, Rebecca Elliott, Nils Muhlert

## Abstract

Repeated overstimulation of the stress response system, caused by exposure to prolonged highly stressful experiences, is thought to affect brain structure, cognitive ability, and mental health. We tested the effects of highly stressful experiences during childhood and adulthood using data from the UK Biobank, a large-scale national health and biomedical study with over 500,000 participants. To do this, we defined four groups with high and low levels of childhood and adulthood stress. We then used T1- and diffusion-weighted MRI data to assess the macrostructure in grey matter and microstructure in white matter of limbic brain regions, commonly associated with the stress response. We also compared executive function and working memory between these groups. Our findings suggest that in females, higher levels of both childhood and adulthood stress were associated with reduced connectivity within the posterior thalamic radiation. High stress in both childhood and adulthood was associated with decreases in both executive function and working memory. Finally, stress across the lifespan was positively associated with the number of diagnosed mental health problems, with a stronger effect in females than in males. Together our findings demonstrate links between stress across the lifespan, brain structure and mental health outcomes that may differ between males and females. Our findings also suggest that exposure to highly stressful life events has a negative impact on cognitive abilities in later life regardless of sex.

## 1. Introduction

Cumulative life stress is the accumulation of repeated exposure to stressful experiences across the lifespan. During childhood, highly stressful events (such as abuse or the loss of a parent) have been shown to impact an individual’s cognitive abilities and both physical and mental health decades later as adults (1,2). Similarly, stressful events during adulthood, such as separation, bereavement or financial difficulties, are amongst the most influential risk factors for coronary heart disease (3), and are also associated with slower processing speed (4). The timing at which stress occurs, either during periods of rapid brain development early in life, or later during periods of relative brain stability, may differentially impact upon brain structure to influence cognitive abilities and health. Here we assess the relative effects of childhood and adulthood stress on brain macro- and micro-structure, cognitive abilities and mental health diagnoses in a very large cohort, using data from the UK Biobank (https://www.ukbiobank.ac.uk).

During childhood, overexposure to stress hormones can suppress immune responses, leaving individuals vulnerable to infection and to disrupted development of the brain (5). For instance, sexual abuse experienced during childhood has been associated with reduced hippocampal volume (6). Additionally, neglect and low socioeconomic status during childhood have been associated with reduced regional brain volumes (7,8), such as the volume of grey matter (9) and white matter (7) in the hippocampus. Reductions in regional brain volumes associated with childhood stress are still detectable much later in life, suggesting a persistent influence of childhood stress on the brain (7). Stress during adulthood is also associated with lower hippocampal volumes (10–13) potentially due to disrupted patterns of neurogenesis (14). The comparative effects of childhood compared to adulthood stress is, however, not yet well understood.

In addition to changes in the volume of brain regions, alterations in microstructure may also be observed in white matter tracts linking these regions. Diffusion MRI can be used to probe the barriers to water diffusion in the brain non-invasively and *in vivo*; it is sensitive to a loss of those barriers, such as through loss of myelin or axonal membranes. High levels of childhood stress are associated with reductions in diffusion metrics in white matter regions including the corpus callosum and uncinate fasciculus (7,15–17). Reduced white matter integrity has been shown in adults who report high levels of life stress (18). These findings suggest that stress, either during child- or adulthood, can impact on the microstructure of the brain.

Cognitive abilities, such as memory, rely on regions of the limbic system such as the hippocampus and amygdala, which coincidentally have a high density of receptors for stress-related hormones (19–22). As such, links between acute stress and worse cognitive performance (including divided attention and working memory) have been established across a wide range of studies (23). Similarly, high levels of perceived stress predict the frequency of everyday cognitive failures (such as forgetting appointments etc), demonstrating the real-world impact of stress on cognition (24). This negative impact on cognition can either occur from residual effects of stress during childhood, where it is associated with worse working memory and higher-order complex functions (25,26), or from stressors during adulthood (4). Changes in these cognitive abilities relating to stress may be underpinned by consequent alterations in limbic structures, such as the hippocampus and amygdala (27).

Mental health outcomes in later life are also linked to stressful experiences in childhood and adulthood (28,29). A history of childhood trauma is associated with smaller hippocampal volumes, as seen in those with stress-related mental health problems including major depression and post-traumatic stress disorder (30–32). In addition to altered regional volumes, major depression has been linked to reduced microstructural integrity in the posterior thalamic radiation (33). This damage or atrophy to microstructure in individuals with major depression has also been linked to stress (34). It therefore seems plausible that stressful experiences across the lifespan impact on both cognition and mental health via their effects on brain structure.

Sex differences in stressor prevalence may also differentially impact upon consequent biological effects. Prevalence rates of extreme stressors, such as sexual and emotional abuse, are much higher in females than males (35,36). Stress hormones, such as cortisol, may also impact males and females differently (37,38). For example, animal models demonstrate that changes in gonadal hormones during ageing, particularly in females, interact with the regulation of genes relevant to stress reactivity (39–42). This increased stress responsivity is accompanied with alterations in limbic brain structures. This provides a route through which sex differences in stress responsivity could impact upon brain structure, and so upon cognition and mental health.

Using the full UK Biobank sample (*n* = 502,520), we first identified four specific categories of individuals based on high or low levels of stress experienced in childhood (CS) and/or adulthood (AS). We examined the relative impacts of stress on brain structure, both through grey matter regional volumes and white matter microstructural metrics in key regions of the limbic system. We predicted that those with high levels of CS and AS would show a reduction in grey matter volume in these limbic regions. We additionally predicted changes in the white matter microstructure between these regions, estimated using diffusion MRI metrics. Finally, we predicted that those with high CS and AS would be more prone to worse cognitive performance and increased numbers of mental health diagnoses.

## 2. Methods

### 2.1. Participants

All participant datasets for this study were obtained from the UK Biobank from data released in February 2020 (https://www.ukbiobank.ac.uk/). The UK Biobank is a large-scale study collecting health and biomedical data from 500,000 generally healthy participants across Great Britain, aged 40-69 years between 2006 and 2010. A subset of these participants were later invited to a second visit to undergo brain MRI. Ethical approval was granted to the UK Biobank by the North West Multi-Centre Research Ethics Committee (REC reference 11/NW/0382) and all participants provided informed consent to participate and for their anonymised data to be used. The current study was conducted under approved UK Biobank application number 49224.

### 2.2. Defining stress groups

As part of the assessment at the UK Biobank, participants completed several online follow-up questionnaires. Before defining stress groups, participants with neurological conditions were removed from the dataset. We then estimated CS or AS using the following questions: CS was assessed using five questions (Field ID 20489-20491) relating to how well-loved and looked after participants were as children, each answered on a five-point rating scale with reverse coding where necessary (see Appendix A). These questions were based on a shortened version of the Childhood Trauma Questionnaire (CTS-5) (43), a modified version of an established and widely used measure of adverse life experiences occurring during childhood (44). These variables have also been used as a measure of childhood adverse life events in previous research using UK Biobank data (45). AS was measured using a similar set of five questions (Field ID 20521-20525) relating to the experience of potentially abusive relationships experienced as an adult, also using a five-point scale (see Appendix B). Scores of total CS and AS were calculated as the sum of each of these five questions respectively. Participants with missing data on these questionnaires were excluded from the analysis (*n* = 147,131).

Once scores for CS and AS had been calculated, groups for high and low CS and AS were defined. Due to a floor effect leading to a severe skew in the number of participants scoring the lowest possible scores for both CS and AS, criteria for low groups was set to scores of 0. As there are no set criteria for defining “high levels” of trauma/stress our *a priori* criteria for high stress was set as 2 standard deviations above the mean (CS = 6.55, AS = 7.05). Using these criteria, we were able to define our four stress groups: 1) low CS & AS (LC/LA), 2) low CS but high AS (LC/HA), 3) high CS but low AS (HC/LA) and 4) high CS & AS (HC/HA).

Not all participants that met the inclusion criteria for one of the four groups had all relevant data for both imaging and cognitive task analyses. Separate samples were therefore created for imaging and cognitive function analyses. Figure 1 shows a flow chart of how group sizes were determined.

**Figure 1.**
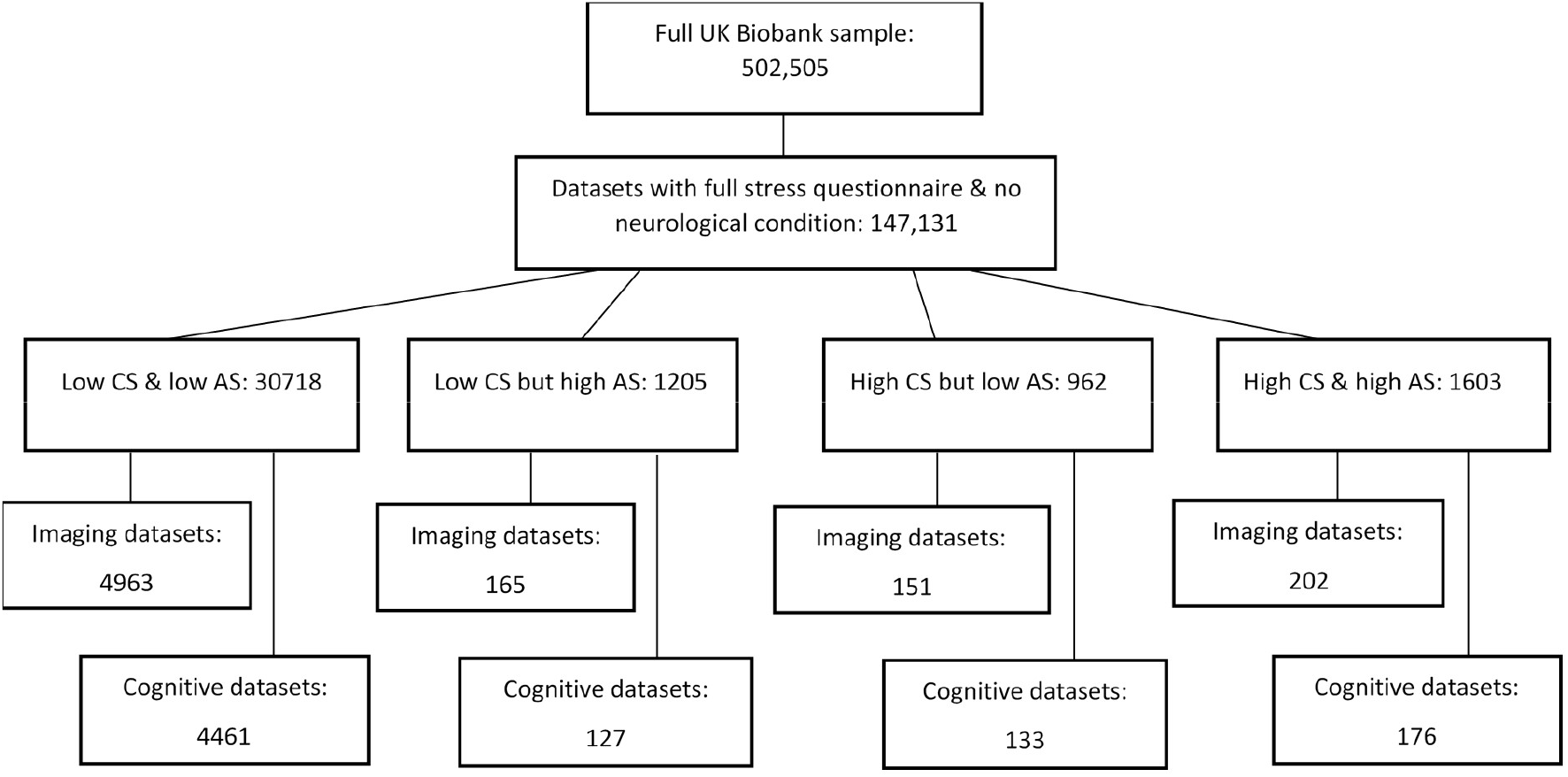
Flow chart demonstrating sample size at each stage of group selection.

### 2.3. Brain image acquisition & processing

Full details of the MRI protocol and processing steps have previously been reported (46,47). The UK Biobank used three dedicated imaging centres, each equipped with identical MR scanners (3.0 T Siemens Skyra, software VD13) using the standard Siemens 32-channel receive head coil. 3D MPRAGE T1-weighted volumes were both pre-processed and analysed by the UK Biobank imaging team using FSL (https://fsl.fmrib.ox.ac.uk/fsl/fslwiki) tools and analysis pipelines adapted from the Human Connectome Project (47–49). The current project takes advantage of the UK Biobank imaging team’s release of analysed imaging data regional statistics, known as Imaging Derived Phenotypes (IDP) (46).

#### 2.3.1. Regional volume analyses

To compare regional volumes, this study used IDP summary values generated by the UK Biobank imaging team (46). After pre-processing these IDPs were created using FSL tools (FIRST) to generate regional grey matter volumes (50). The available limbic structures analysed in the current study were left and right hippocampus, amygdala and thalamus. These regions are shown in figure 2.

**Figure 2.**
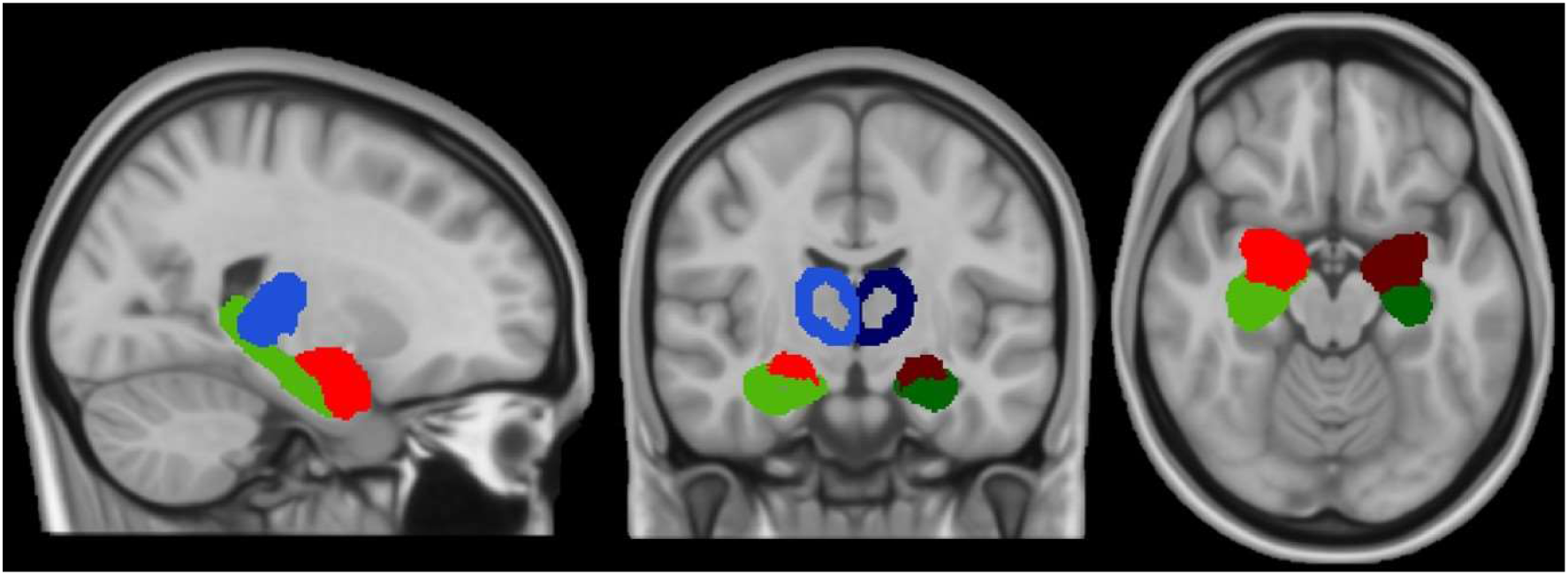
Regional volume maps for amygdala (red), hippocampus (green) and thalamus (blue) in the right (lighter shades) and left (darker shades) hemispheres.

#### 2.3.2. Microstructural analyses

Diffusion MRI metrics were also pre-processed and analysed by the UK Biobank imaging team and the current study used the IDP summary measures generated for all analyses in this study (46). These IDPs include DTI and NODDI (Zhang, Schneider, Wheeler-Kingshott, & Alexander, 2012) measures in major white matter tracts. DTI fitting provides values for mean diffusivity (MD), fractional anisotropy (FA), diffusion tensor mode (MO) and eigenvalues (L1, L2 & L3), whereas the NODDI model provides neurite orientation dispersion (OD) in addition to intra-cellular volume fraction (ICVF, i.e. neurite density) and isotropic volume fraction (ISOVF). All diffusion metrics were analysed between groups in white matter regions: left and right posterior thalamic radiation and cingulum of the hippocampus (see figure 3). No diffusion data were available for the amygdala.

**Figure 3.**
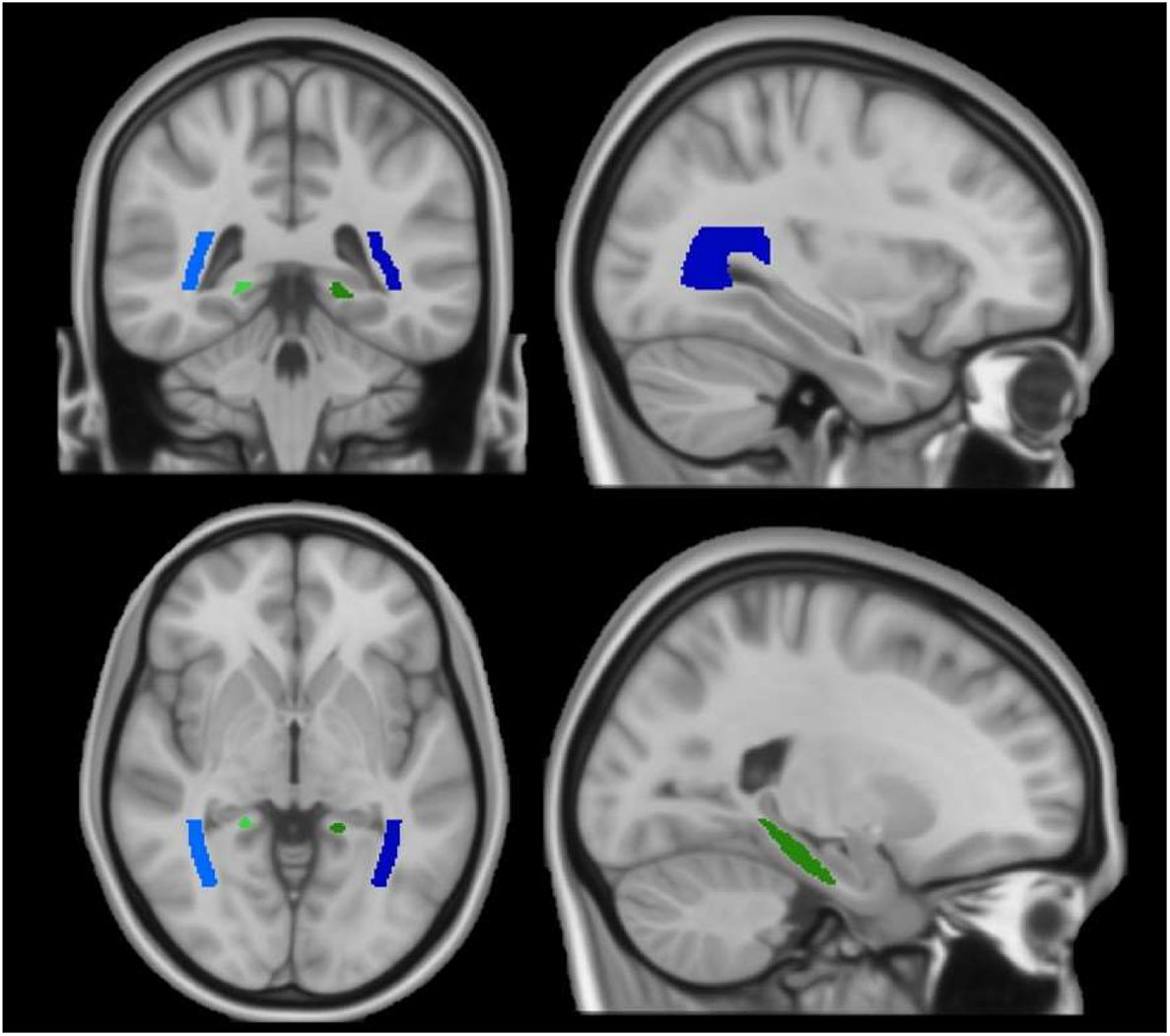
White matter region maps for cingulum of the hippocampus (green) and posterior thalamic radiation (blue) in the right (lighter shades) and left (darker shades) hemispheres.

### 2.4. Cognitive function tasks

All cognitive function tasks were performed on a touchscreen during each participant’s assessment centre visits. Scores for five of these tasks were analysed in this study: numeric memory, prospective memory, symbol digit substitution, trail making and word production. The procedures for these tasks have previously been reported (52). These tasks have been shown to have good concurrent validity and test-retest reliability (53).

### 2.5. Mental health reporting

As part of the online follow-up questionnaires, participants were asked to report if they had ever had mental health problems diagnosed by a professional (Data Field 20544). Summative scores were calculated to determine how many diagnosed mental health problems each participant had experienced.

### 2.6. Statistical analysis

The handling of raw UK Biobank data, creation of sub samples and groups and the statistical analysis for all comparisons were all conducted using R (54). Individuals reporting neurological conditions or with missing data for measures of interest in relevant sub samples were excluded. Kruskal Wallis and chi-squared tests were used to compare groups’ ages and sex ratios, respectively.

To compare group brain volumes, diffusion metrics and cognitive scores, between-subjects ANOVA tests were used. However, as it was likely that other factors including age and sex may have affected the data, these factors were added as covariates, resulting in the use of between-groups ANCOVA tests. As multiple regions were compared in the imaging analyses, and multiple tasks used in the cognitive analyses, false discovery rate (FDR) multiple comparisons corrections were used to adjust significance values (*p*-values). Only comparisons surviving multiple corrections were reported as significant. *Post hoc* pairwise comparisons were used to establish between which stress group any significant differences occurred. *Post hoc* Pearson’s r correlations were also used to explore potential relationships between stress and brain imaging measures in male and female samples separately across the full sample with appropriate imaging data. Age was also be used as a covariate within these correlations.

Due to missing data, cognitive comparisons were only possible for symbol digit substitution and trail making (both numeric and alphanumeric) tasks. These comparisons were conducted using the same ANCOVA models as the brain imaging data, with childhood and adulthood stress as factors and adjusting for age and sex. *Post hoc* pairwise comparisons were used to determine group differences. Pearson’s r correlations within the wider sample (with appropriate data) were performed to assess any relationship between CS, AS and these other cognitive function tasks. Finally, correlations were also used to identify any relationships between the wider UK Biobank sample and mental health diagnoses. Fischer r-to-z transforms may be used to compare the strength of relevant correlations.

## 3. Results

### 3.1. Group differences: Age & Sex

Groups differed significantly in age (Kruskal Wallis: imaging [*χ*^*2*^(3)= 31.9, *p* < .001] and cognitive data [*χ*^*2*^ (3)= 30.96, *p* < .001]) and sex (Chi-squared: imaging [*χ*^*2*^ (3)= 134.2, *p* < .001] and cognitive data [*χ*^*2*^ (3)= 125.4, *p* < .001]; Table 1).

**Table 1.** Number of participants, mean ages (with standard deviation in parentheses) and sex ratios for each stress group across both imaging and cognitive datasets. Sex ratio is female to male.

|  | Brain imaging |  |  | Cognitive Data |  |  |
| --- | --- | --- | --- | --- | --- | --- |
|  | *N* | Age | Sex | *N* | Age | Sex |
| Group 1<br>(Low CS & Low AS) | 4963 | 55.16<br>(7.16) | 1.07 | 4461 | 55.16<br>(7.29) | 1.04 |
| Group 2<br>(Low CS & High AS) | 165 | 53.54<br>(7.30) | 3.85 | 127 | 52.92<br>(7.44) | 3.88 |
| Group 3<br>(High CS & Low AS) | 151 | 54.07<br>(7.93) | 1.7 | 133 | 53.37<br>(7.71) | 1.89 |
| Group 4<br>(High CS & High AS) | 202 | 52.66<br>(7.24) | 5.5 | 176 | 52.91<br>(7.58) | 5.77 |

### 3.2. Impact of stress on brain volume

Group differences in grey matter volume of the limbic structure regions, left and right hippocampus, amygdala and thalamus, scaled to account for head size differences, were assessed using between-subject ANCOVA tests.

No significant volume differences were seen between groups for any of the six regions of interest. As expected, both age and sex had large significant effects on both hippocampal and thalamic volume (for each analysis, *p* < .0001), but not amygdala volume.

### 3.3. Impact of stress on brain microstructure

Age was a significant covariate for all diffusion metrics in the posterior thalamic radiation. Sex was a significant covariate in bilateral FA, MD, L1 & ICVF, and right MO but not for ISOVF or OD.

Significant group effects were seen in the right and left posterior thalamic radiation for FA (*F*(3, 5475) = 4.47, *p* = .014, 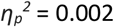 & *F*(3,5475) = 4.02, *p* = .014, 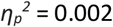 and left posterior thalamic radiation for ISOVF (*F*(3, 5475) = 5.42, *p* = .004, 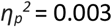 Data distributions for FA and ISOVF in bilateral posterior thalamic radiation can be seen in Figure 4.

**Figure 4.**
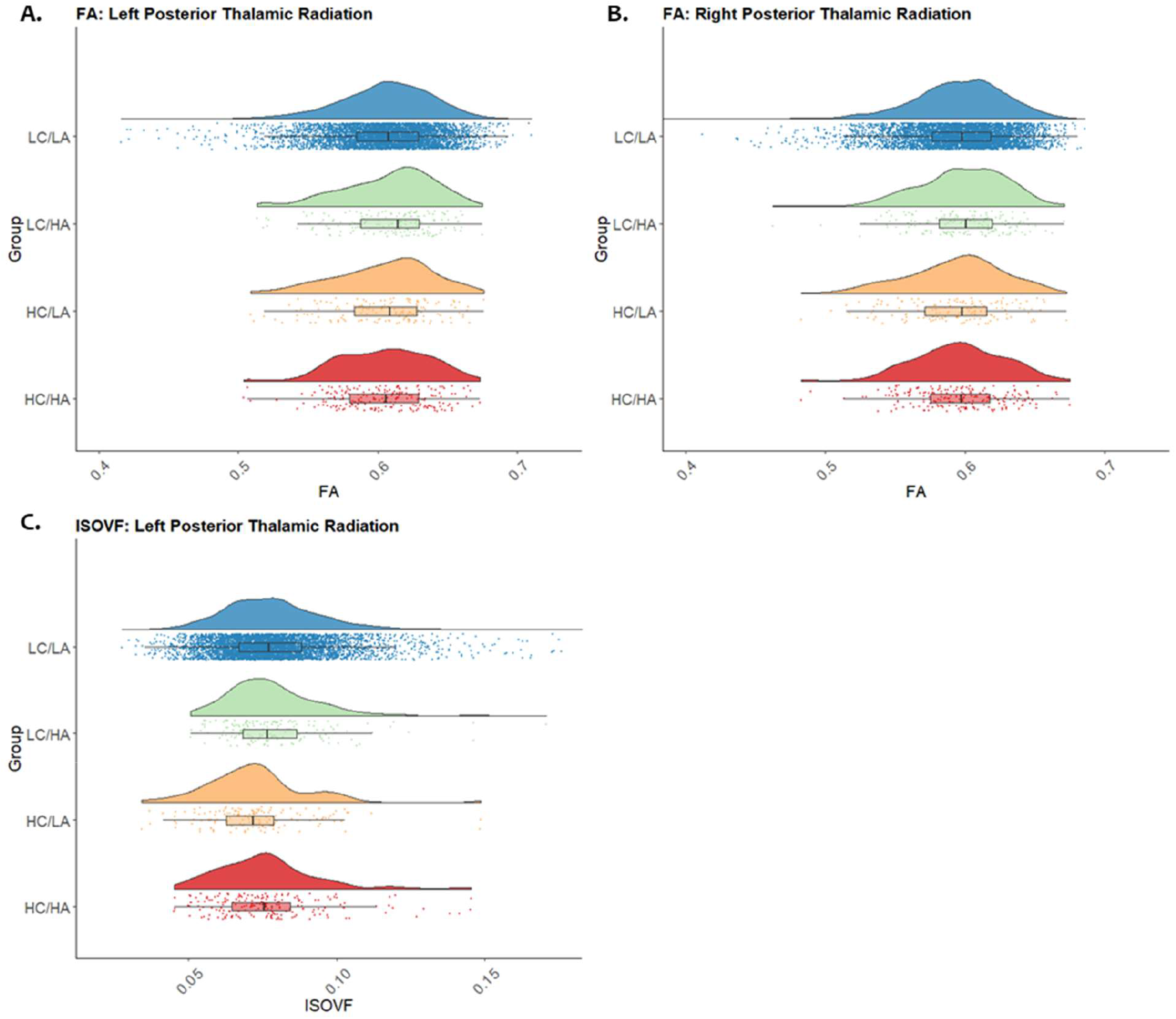
Raincloud plots representing data distributions in each of the four stress groups for values of FA in the left (A) and right (B) posterior thalamic radiation & ISOVF in the left (C) posterior thalamic radiation between each stress group. Box plots represent median and interquartile ranges of these values.

*Post hoc* pairwise comparisons revealed reduced FA in the right and left posterior thalamic radiation for HC/HA compared to the LC/LA group (right: *p* = .018, 0.008% reduction in mean FA, Left: *p* = .009, 0.11% reduction in mean FA). The HC/LA group also showed significantly lower left posterior thalamic radiation ISOVF compared to LC/LA (*p* = .0005, 7.88% reduction mean ISOVF), LC/HA (*p* = .003, 8.24% reduction in mean ISOVF), and HC/HA (*p* = .035, 5.73% reduction in mean ISOVF) groups.

No significant effects of group were seen for any diffusion metric in the left or right cingulum of the hippocampus. Significant effects of age as a covariate were seen for all diffusion metrics in the cingulum of the hippocampus, except for MO on the right side. Additionally, sex was a significant covariate in left and right cingulum of the hippocampus for FA, MO, ICVF, L2 and ISOVF. For MD, L1 and L3, sex was only a significant covariate in the right and OD in the left.

### 3.4. Sex differences in the brain

Given the significant effects of sex in the above analyses, *post hoc* examinations of the effects of stress on brain structure in males and females separately. For all male (N = 11,290) and female (N = 13,285) participants with sufficient stress and imaging data, partial correlations (with age as a covariate) were run for brain volumes and diffusion metrics in the associated regions of interest. FDR corrections for multiple comparisons were used. In males, adulthood stress correlated with FA and OD in the left posterior thalamic radiation (*r* = -0.033, *p* = .003 & *r* = 0.026, *p* = .026) and with ISOVF in the left (*r* = -0.026, *p* = .03) and right (*r* = -0.03, *p* = .03) cingulum of the hippocampus. Childhood stress was significantly correlated with FA in the right posterior thalamic radiation (*r* = - 0.026, *p* = .02).

In females we observed significant correlations between CS, AS and several volume and diffusion measures (Table 2). Fisher r-to-z transforms were used to compare CS and AS correlations with MD in the cingulum of the hippocampus and posterior thalamic radiation. In the right posterior thalamic radiation, CS showed a significantly stronger correlation with MD than AS (*z* = 1.73, *p* = .042).

**Table 2.**
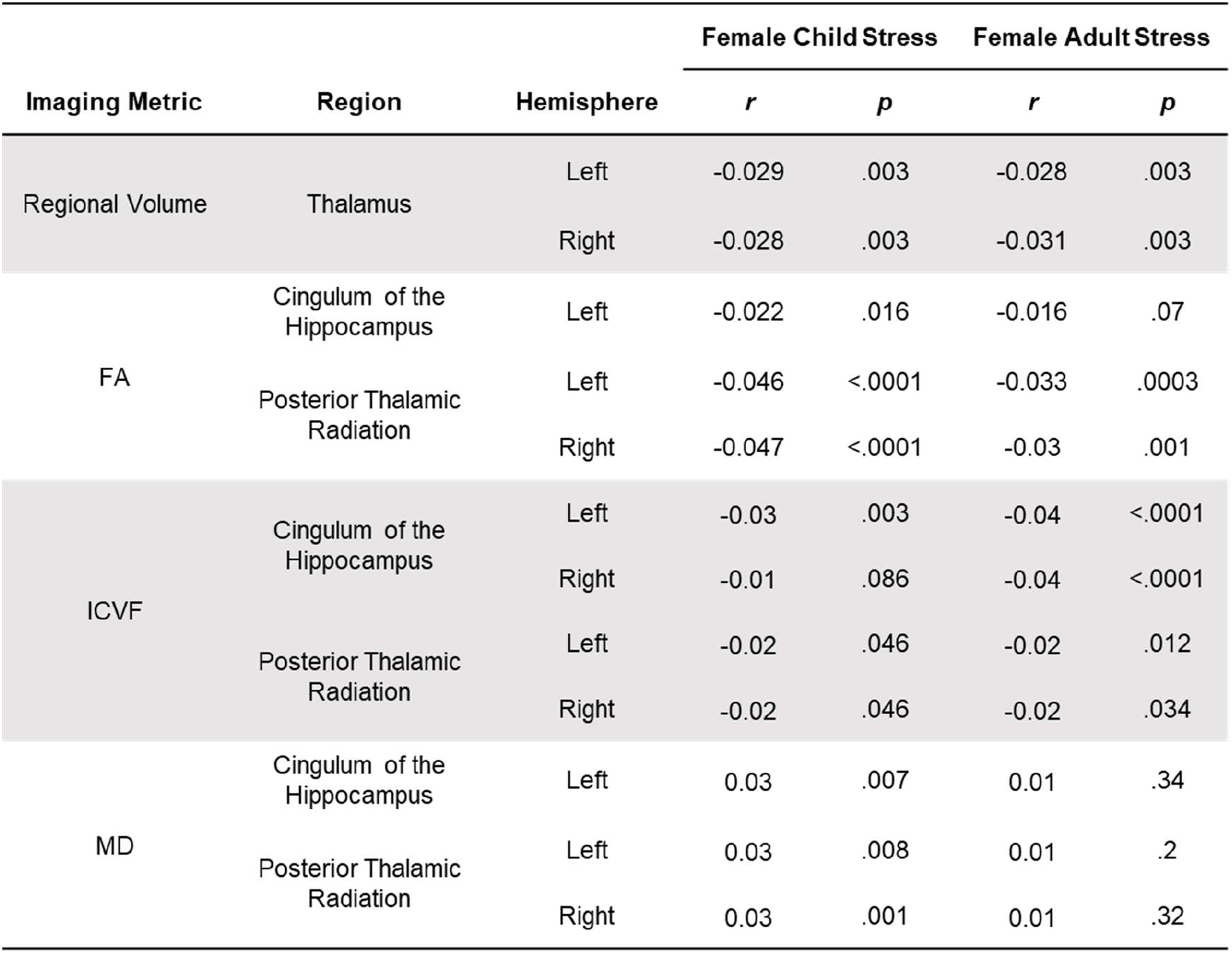
Correlations for significant brain metrics with child and adult stress in female participants only.

### 3.5. Impact of childhood and adulthood stress on cognitive function

ANCOVA tests were used to identify group differences in performance on all three cognitive tasks using FDR to correct for multiple comparisons (Figure 5). Symbol digit substitution performance was linked to both group (*F*[3, 4891] = 10.86, *p* < .0001, 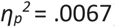). *Post hoc* pairwise comparisons revealed worse performance in HC/HA than LC/LA (*p* < .0001, 6.67% reduction in mean score) and HC/LA (*p* = .005, 8.14% reduction in mean score) groups.

**Figure 5.**
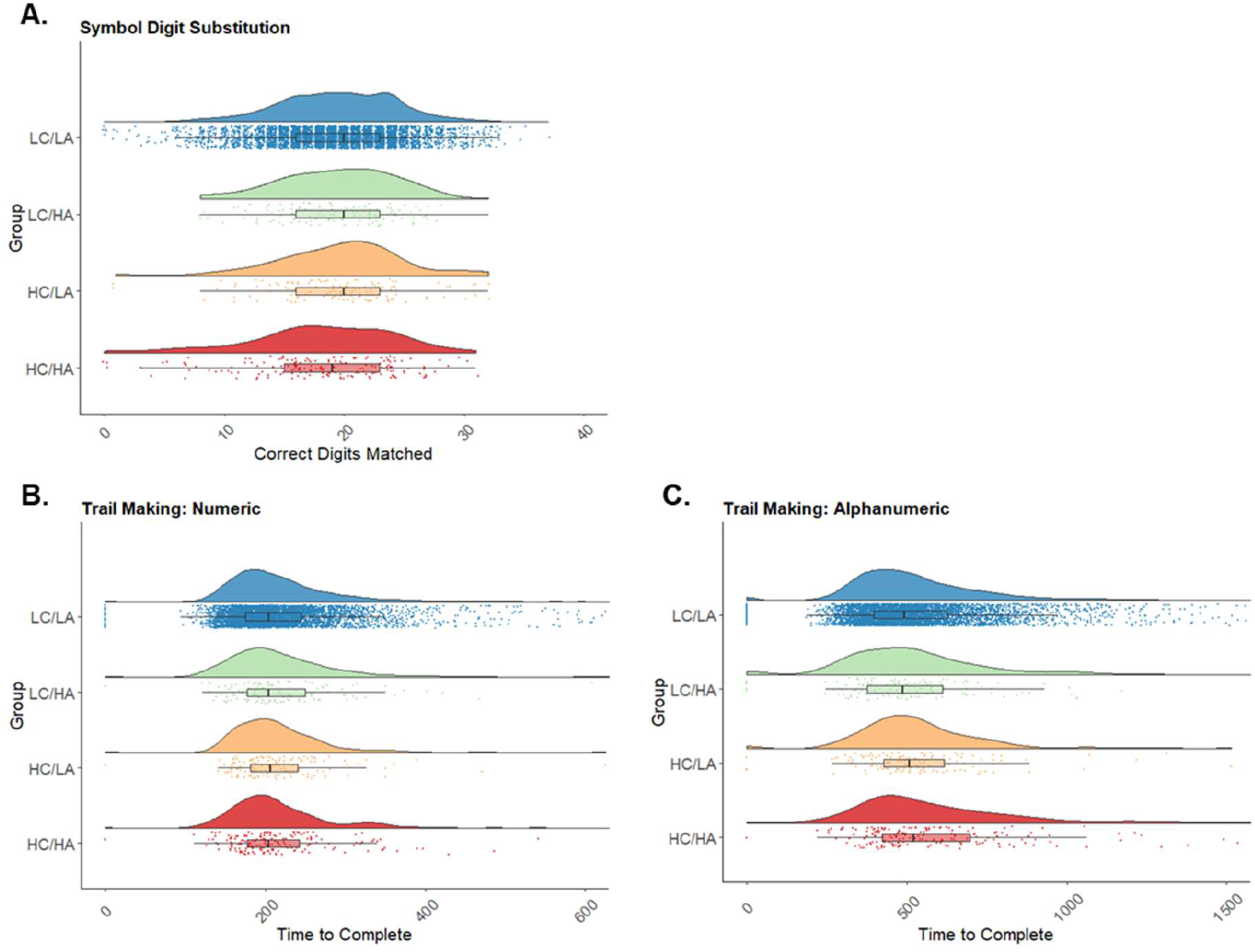
Raincloud plots representing data distributions in each of the four stress groups for (A) correct digits matches for symbol digit substitution task, (B) duration to complete the numeric trail making task (C) duration to complete the alphanumeric trail making task. Box plots represent median and interquartile ranges of these values.

Numeric trail making performance also revealed a significant effect of group (*F*(3, 4891)= 3.75, *p* = .01, 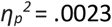), however *post hoc* analyses show only a non-significant trend for worse performance in the HC/HA than the LC/LA group (*p* = .06, 2.29% increase in mean time to complete). In contrast, alphanumeric trail making performance also showed group differences (*F*(3,4891) = 6.47, *p* = .0003, 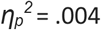), but with significantly worse performance in the HC/HA than LC/LA (*p* < .001, 10.28% increase in mean time to complete) and LC/HA groups (*p* = .02, 15.3% increase in mean time to complete).

In the whole sample we found significant correlations between levels of CS and AS and performance on each of the cognitive tasks (Table 3), except for word production scores.

**Table 3.** Sample size, correlation and significance values for child and adult stress with cognitive tasks.

| Cognitive task | N | Child Stress |  | Adult Stress |  |
| --- | --- | --- | --- | --- | --- |
|  |  | r | p | r | p |
| Numeric Memory | 15925 | -0.069 | <.0001 | -0.1 | <.0001 |
| Symbol Digit Substitution | 20434 | -0.074 | <.0001 | -0.08 | <.0001 |
| Numeric Trail Making | 20596 | 0.040 | <.0001 | 0.049 | <.0001 |
| Alphanumeric Trail Making | 20596 | 0.054 | <.0001 | 0.067 | <.0001 |
| Word Production | 853 | -0.052 | .133 | -0.072 | .037 |

### 3.6. Impact of stress on mental health

Correlations assessed the relationship between CS, AS and mental health outcomes (*N* = 146,206). We found significant associations between the number of mental health issues reported and both CS (*r* = .22, *p* <. 001) and AS (*r* = .21, *p* > .001) (Figure 6).

**Figure 6.**
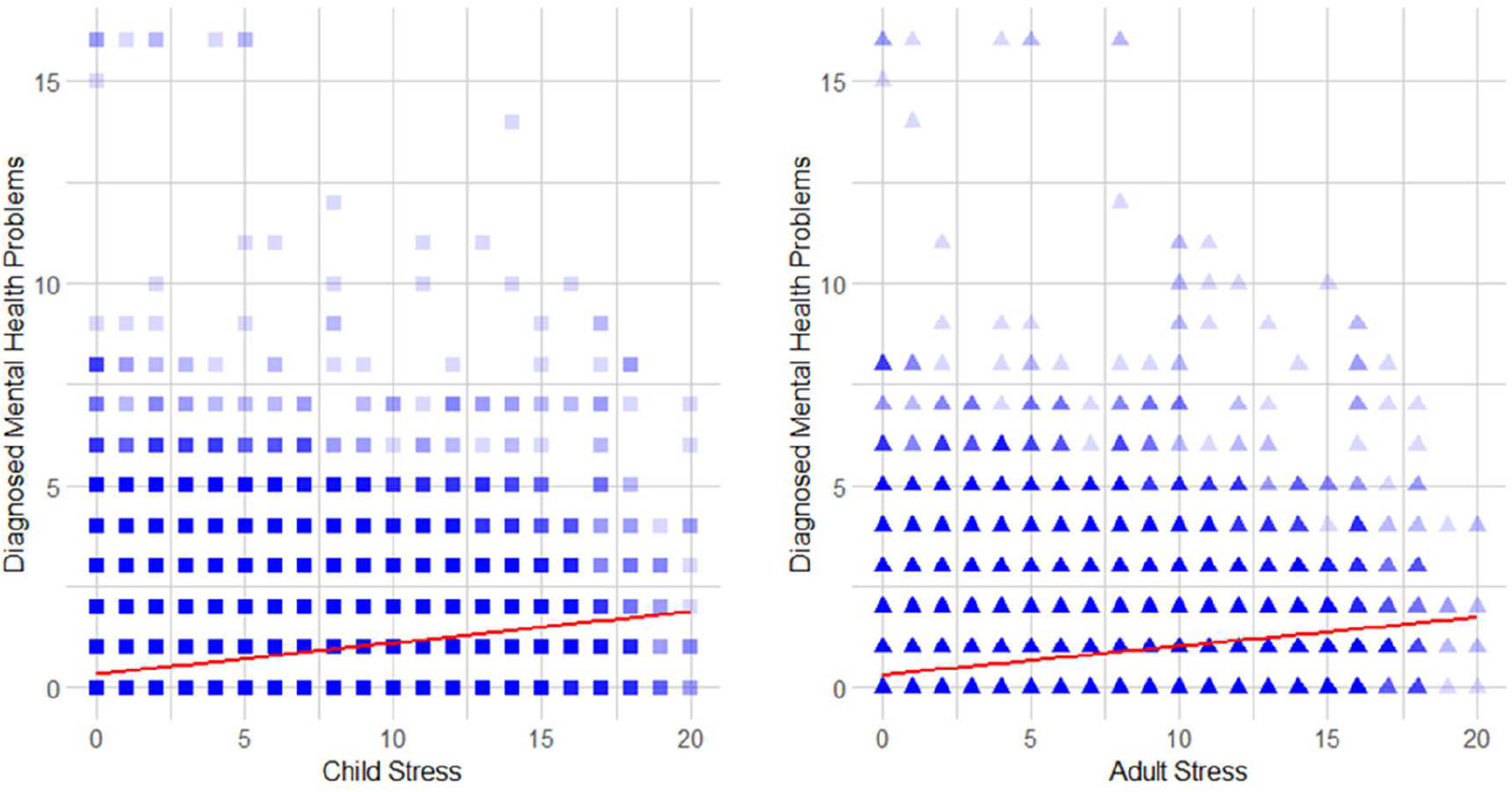
Correlation scatter plots for child stress (left) and adult stress (right) with the number of mental health diagnoses participants have received, with the linear trend line in red. Darker colour represents more participants scoring those values.

We also examined correlations between mental health outcomes and stress for males (*N* = 64,453) and females (*N* = 81,755) separately. In males, both CS and AS correlated with mental health outcomes (*r* = .18, *p* <.001, & *r* = .15, *p* < .001, respectively). In females, strong correlations were also seen between mental health outcomes, CS and AS (*r* = .27, *p* < .001, & *r* = .22, *p* < .001, respectively). Using a Fisher r-to-z transformation, it was shown that these correlations were significantly stronger for females both for CS *(z* = 18.01, *p* < .001) and AS (*z* = 13.77, *p* < .001) than males.

## 4. Discussion

We examined the relationship between reported childhood and adulthood stress and measures of brain macro- and microstructure, as well as cognitive and mental health outcomes. We demonstrate that stress at both stages of the lifespan is linked to microstructural changes within the bilateral posterior thalamic radiation. Changes in ISOVF may indicate infiltration of CSF within a region, a potential marker of tissue breakdown.

*Post hoc* analyses revealed substantial differences in the effects of stress on the brains of male and female participants. In females there were significant correlations between stress (both child and adult) and microstructural measures (FA and ICVF) in the posterior thalamic radiation and cingulum of the hippocampus. The effect sizes for these correlations were, however, very small. Additionally, associations were shown between increased MD and childhood stress in females alone, potentially suggesting specific effects of childhood stress on the female brain. These findings are in line with previous literature, which reports reduced FA and increased MD in females only (55,56). We also found that females who had experienced more childhood and adulthood stress showed smaller thalamic volumes, indicating both macro- and microstructural effects on thalamic brain structure and outputs. It has previously been argued that stressful experiences in females have a greater detrimental effect on brain structure than in males (37,38). This may occur through interactions between stress-related hormones and oestrogen. This is supported by evidence that treatment with oestrogen is associated with decreased FA and increased MD (55). How oestrogen interacts with cortisol to affect brain microstructure however needs careful study.

Cognitive performance was worse in those with a history of both childhood and adulthood stress. Unlike brain structure, these effects were not different in males and females. These findings are in line with previous research regarding both acute and life stress effects of stress on cognition (24,57). More specifically, stress and particularly adulthood stress is associated with worse performance on alphanumeric trail making tasks. This finding links with previous evidence suggesting that stress during adulthood leads to specific impairments in processing speed, a key factor within alphanumeric trail making tasks (4,58). Higher stress throughout the lifespan, but particularly within childhood, was associated with worse symbol digit substitution performance. When analysing all available participants from the UK Biobank with stress data, we found correlations between these cognitive tasks and both childhood and adulthood stress. The use of converging tasks that isolate and examine individual cognitive processes can help to confirm whether, and understand why, these processes would be vulnerable to stress at different life stages.

As expected, we show clear evidence linking increased levels of both child- and adult-hood stress with increased numbers of mental health diagnoses. Although this effect is clear in both males and females, it is particularly striking, and indeed significantly greater, in females. In line with this, it has previously been reported that females show higher frequencies of stress-related affective and mental health disorders (39,59–61). In the brain, microstructural alterations within the posterior thalamic radiation have previously been associated with major depression (33). As our findings demonstrate alterations in posterior thalamic radiation, stress may be a key causal factor underpinning the relationship between brain structures and mental health.

Our study is not without limitations. Only a subsection of UK Biobank participants had completed neuroimaging (*N* = 5481) and cognitive (*N* = 4897) measures. While we were not testing the same participants in each analysis, the considerable sample sizes should allow for robust conclusions to be drawn. There were fewer participants with high levels of child- and adulthood stress than those with low levels of stress throughout the lifespan. This may reflect the low frequency of these extreme adverse events but may also reflect the potential lack of representation within the biobank of individuals from deprived communities where these stressors may be more prevalent. Similarly, there were comparably greater numbers of females in the high stress groups, reflecting the greater likelihood of experiencing abuse and control in women (35,36). To account for this, we used sex as a covariate where appropriate and analysed sex-differences in our sample.

Additionally, the comparison of macro- and microstructural measures come from different areas (macrostructure in grey matter and microstructure in closely related white matter tracts) making it difficult to compare the two directly as any differences observed may also be due to the difference in location. Future research can directly compare macro- and microstructural changes in the same regions in order to better draw comparisons regarding the relative impact of stress on these measures. A further limitation concerns our definitions of childhood and adulthood stress. The UK Biobank conducted no specific measure of child and adult life stress. The measure of childhood stress was based on the CTS-5 (43), however this may not reflect all possible experiences of stress in childhood. We also used the available data to select comparable questions relating to adult life experiences.

In the present study, we have demonstrated that high levels of stress at different points in the lifespan show specific associations with decreased performance on different cognitive tasks. In females, we found subtle alterations in brain microstructure associated with higher levels of stress, particularly in the posterior thalamic radiation. Finally, we demonstrate that stress during both childhood and adulthood is associated with an increased incidence of diagnosed mental health outcomes. These findings shed light on how stressful experiences throughout the lifespan may impact upon the brain, cognition, and mental health outcomes.

## Data Availability

The data for this project was provided by the UK Biobank (application number 49224)

## Acknowledgements

This work was funded by the BBSRC as part of a DTP studentship. The data for this project was provided by the UK Biobank (application number 49224).

## 6. Appendices

### Appendix A

Items used to create child stress scores. Category 145: Online follow up-Mental health-Traumatic events. Field IDs, child stress statement, use of reverse coding and number of each response to the question.

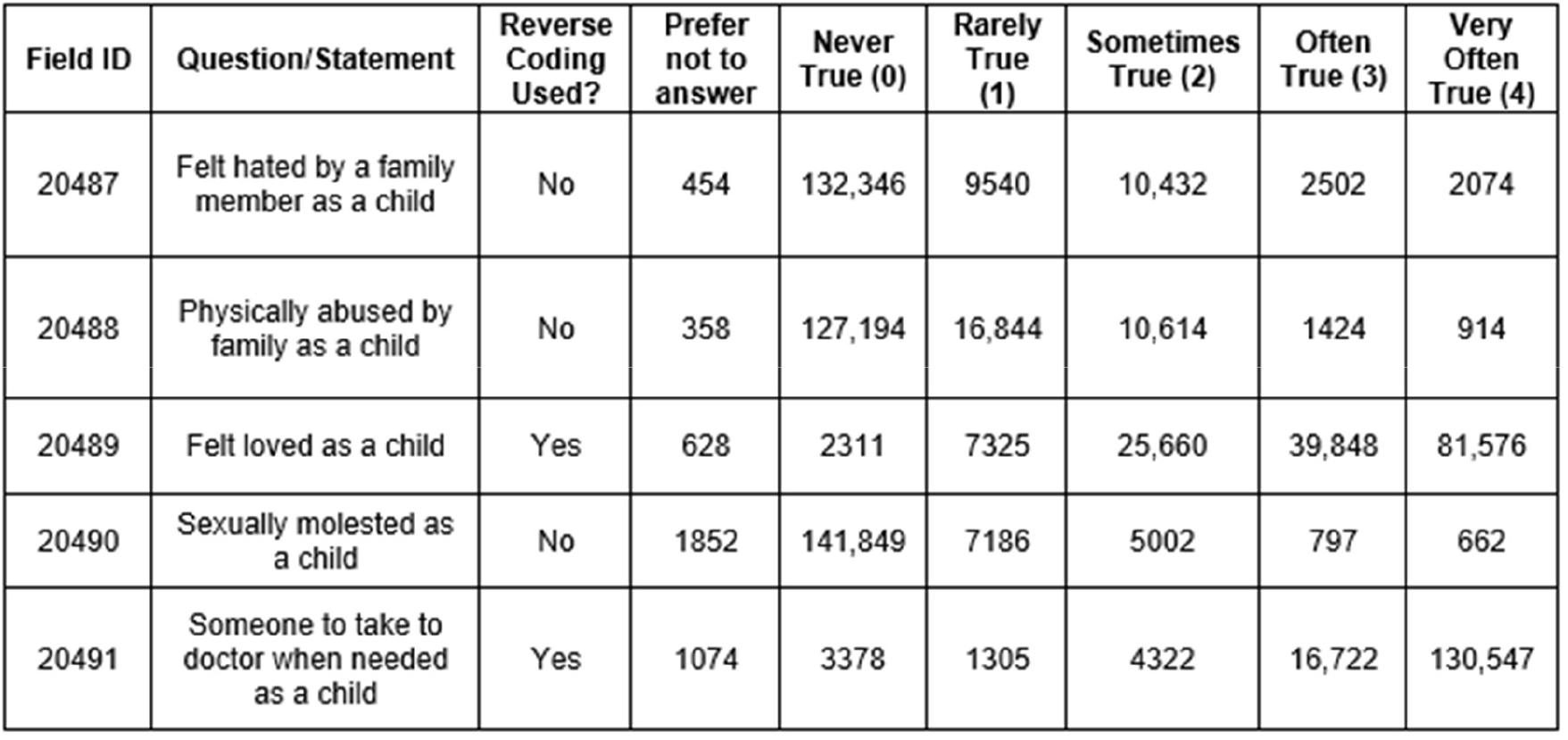

| Field ID | Question/Statement | Reverse Coding Used? | Prefer not to answer | Never True (0) | Rarely True (1) | Sometimes True (2) | Often True (3) | Very Often True (4) |
| --- | --- | --- | --- | --- | --- | --- | --- | --- |
| 20487 | Felt hated by a family member as a child | No | 454 | 132,346 | 9540 | 10,432 | 2502 | 2074 |
| 20488 | Physically abused by family as a child | No | 358 | 127,194 | 16,844 | 10,614 | 1424 | 914 |
| 20489 | Felt loved as a child | Yes | 628 | 2311 | 7325 | 25,660 | 39,848 | 81,576 |
| 20490 | Sexually molested as a child | No | 1852 | 141,849 | 7186 | 5002 | 797 | 662 |
| 20491 | Someone to take to doctor when needed as a child | Yes | 1074 | 3378 | 1305 | 4322 | 16,722 | 130,547 |

### Appendix B

Two sets of statements and response keys used to create adult stress scores. Category 145: Online follow up-Mental health-Traumatic events. Field IDs, adult stress statement, use of reverse coding and number of each response to the question.

| Field ID | Question/Statement | Reverse Coding Used? | Prefer not to answer | Never True (0) | Rarely True (1) | Sometimes True (2) | Often True (3) | Very Often True (4) |
| --- | --- | --- | --- | --- | --- | --- | --- | --- |
| 20521 | Belittlement by partner or ex-partner as an adult | No | 485 | 119,234 | 14,964 | 15,724 | 3669 | 3272 |
| 20522 | Been in a confiding relationship as an adult | Yes | 4127 | 14,051 | 7401 | 27,692 | 26,848 | 77,229 |
| 20523 | Physical violence by a partner or ex-partner as an adult | No | 491 | 136,776 | 9877 | 7408 | 1291 | 1505 |
| 20524 | Sexual interference by a partner or ex-partner without consent as an adult | No | 515 | 147,762 | 5028 | 3035 | 478 | 530 |
| 20525 | Able to pay rent/mortgage as an adult | Yes | 2448 | 5628 | 1109 | 4031 | 11969 | 132,163 |

